# DEVELOPMENT OF A COMPLEX INTERVENTION PACKAGE FOR DENGUE PREVENTION

**DOI:** 10.1101/2022.06.16.22274559

**Authors:** R. M. Nayani Umesha Rajapaksha, Chrishantha Abeysena, Aindralal Balasuriya, Nimalka Pannila Hetti, Ajith Alagiyawanna, Suranga Manilgama

**Affiliations:** Ministry of Health, Colombo, Sri Lanka; Department of Community Medicine, Faculty of Medicine, Ragama, University of Kelaniya, Sri Lanka; General Sir John Kotelawala Defence University, Ratmalana, Sri Lanka; National Dengue Control Unit, Ministry of Health, Colombo, Sri Lanka; Health Promotion Bureau, Ministry of Health, Colombo, Sri Lanka; Teaching Hospital Kurunegala, Sri Lanka

**Keywords:** “dengue”, “communication”, “behaviour”, “waste management”, “COMBI”

## Abstract

**Introduction:** Complex interventions are widely used in public health practices. Communication for Behavioural Impact (COMBI) is an effective method in planning for dengue control. The aim was to develop an intervention package to change behaviour to prevent dengue in a dengue endemic area of Sri Lanka.

**Methods:** Development of the intervention package was formulated according to two-phases including ‘Theoretical phase’ and ‘Modelling phase’ using a framework. World Health Organization 10 key steps in planning COMBI were followed in order to develop the present intervention package. A Situational-Market-Analysis-for-Communication-Keys (SMACK) was conducted in highest dengue endemic area in Kurunegala district to identify the Specific Behavioural Objectives (SBOO), which was conducted using mixed methodological approach followed by other steps of COMBI planning.

**Results:** The overall goal was to decrease morbidity and mortality due to dengue illness. The SBOO for the plan were to improve the proper waste management practices according to the ‘3R concept’ (Reduce-Reuse-Re-cycling) and to improve the dengue prevention practices by 30 minutes of weekly cleaning. The strategies were to conduct a community empowerment program to improve waste management and weekly practices on dengue prevention by conducting administrative mobilization and public-relationship, public-advocacy, community mobilization, personal selling, advertising, and point of service promotion. The plan was finalized after taking expert opinion with the participation of stakeholders by the management team.

**Conclusion:** Developing a COMBI plan for an area after the identification of SBOO would be feasible to implement in order to empower the community to prevent dengue and improve community health services.

## Introduction

Dengue is a major health problem affecting mainly the tropical countries. Many strategies have been employed to tackle the outbreaks, but the key components of outbreak response are household and community level behavioural and social interventions. Moreover, leadership and planning for sustainable community participation, transfer of technical knowledge and skills in planning, and measures to ensure sustainability at each level are identified as key issues in dengue prevention. However, behavioural change is important in effective implementation of control activities to prevent and control dengue. Many community-based dengue control programmes focus only to improve knowledge and to increase awareness assuming that changing behaviours,[1].

Complex interventions are widely used in public health practices. Main three stages can be identified in the development of complex intervention, [2-4]. Moreover, systematically used theory to develop the intervention, fully description of the intervention will help to implement properly for the purposes of the evaluation. Importantly, the developed intervention needs to be replicated by others. The behavioural interventions would be focused on identifying key risk reduction actions at the household and community level to minimize negative health consequences. Moreover, application of multiple approaches is essential to be included to achieve the success to ensure community-located outbreak prevention through achieving specific behavioural results,[5]. A systematic review revealed that community mobilization programmes are effective interventions in reducing *Aedes aegypti* entomological indices,[6]. Notably, multi-stakeholder partnership leads to achieve successful outcomes of the interventions. Furthermore, the importance of incorporating health education and capacity building for the development of the programmes to prevent communicable disease is highlighted,[7]. Therefore, there is a need to empower the community through multi-stakeholder participation to achieve sustainable solution for dengue prevention and all steps to be taken to minimize the dengue burden by changing behaviour of the community members. Importantly, systematic development of a complex intervention with minimum risk of bias is currently needed and it needs to be culturally and geographically suited.

Communication for Behavioural Impact (COMBI) is defined as a methodological process which blends strategically blends a variety of communication interventions intended to engage individuals and families in adopting and maintaining healthy behaviours and maintaining those behaviours. It uses a managerial view to plan social mobilization and communication for behavioural impact on public health. The importance of social mobilization for prevention and control of dengue has gathered pace internationally in recent years. Further, COMBI planning offers a comprehensive and innovative managerial insight to planning social mobilization and communication for behavioural impact which is intended for programme managers in integrating interventions to effectively manage dengue. Moreover, to ensure sustainable dengue prevention, the COMBI plans should address the various at-risk populations and it needs to be adopted culturally,[8].

There is a lack paucity of studies with scientifically developed intervention packages for dengue prevention. In this backdrop, to prevent the occurrence of dengue outbreaks, the behaviour of the community needs to be changed. Thus, the development of an intervention for the prevention of dengue based on evidence is a timely need. Importantly, the results of this study would enable responsible authorities to strengthen control strategies to improve dengue prevention activities and it will generate a greater interest to develop capacities for dengue management and would enable them to take action to minimize the impact due to disease burden. The aim was to develop a complex intervention package to change the behaviour of the householders for dengue prevention in Kurunegala district, Sri Lanka.

## Methods

A complex intervention process was formulated according to 2008 guide and modified with the revised framework of ‘Developing and Evaluating Complex Interventions: New Guidence’,[2-4]. This framework provides a guideline for using a stepped approach which separates different elements and using the probable active component in developing the intervention. The development of the intervention package was formulated according to the two phases including ‘Theoretical phase’ and ‘Modelling phase’.

Theoretical phase is to identify evidence base and appropriate theory to gather relevant information to find out the effective intervention packages for dengue prevention. Thus, the facts for the theoretical phase were identified from various sources, including existing evidence, expert opinion, and community need assessment. The modelling phase consists of modelling process and outcomes assessment. The model of the present study was the COMBI theory to empower the community to change the behaviour and ensure sustainable dengue control. The World Health Organization (WHO) 10 key steps in planning COMBI strategies were followed in order to develop the present intervention package,[1,9,10]. Accordingly, a Situational Market Analysis for Communication Keys (SMACK) was conducted in highest dengue endemic area in Kurunegala district (Kurunegala MOH area) to identify the Specific Behavioural Objectives (SBOO) for the plan. The SBOO were finalized using the results of the SMACK which was conducted using mixed methodological approach. Thereafter, the intervention package was finalized by modified Delphi method with expert opinion from specialists in public health from the National Dengue Control Unit (NDCU), Health Promotion Bureau (HPB), Provincial level, general medicine and consensus of the grass-root level experts and an advisor of the international COMBI institute. The developed plan was presented to the panel of expert and finalized the SBOO, strategies, activities for implementation of the intervention and monitoring of the intervention. The management team prepared the scheduling, budget plan with the guidance of an economist, mobilization for resources and activity planning for the implementation of the developed intervention.

## Results

### Theoretical phase

There are different types of behavioural theories for health education, health promotion and behaviour change. Of them, Health Belief Model (HBM),[11,12], Seven Doors Model for Behaviour Change,[13,14], the Community Capacity Building Model for Sustainable Dengue Problem Solution (CCB-SDPS),[15-18], and Communication for Behavioural Impact (COMBI) planning,[1,9,19] were the widely used behavioural changed models for dengue prevention. After considering the effectiveness of the already conducted studies, planning abilities, implementation, and feasibility issues with the consensus of the expert panel, this intervention was formulated by using the COMBI theory. According to the literature review, the components of effective intervention packages were building partnership with multi-stakeholders; waste management at households; improving garbage collection; organization; entomological risk surveillance; capacity building at grass root level; stakeholder meetings and formulation of local steering committee; community working group formation and ensuring intra-sectoral coordination. The major factors associated with sustainable prevention of dengue are adequate knowledge of dengue and dengue prevention, positive attitudes towards the dengue prevention, adequate dengue prevention practices among the community members, adequate health seeking behaviour and high-risk perception on dengue, adequate dengue prevention behaviours, adequate community capacity for sustainable dengue prevention and adequate management of wet containers to prevent vector proliferation. The identified theories and components were used to develop the appropriate model. According to the studies, knowledge, attitudes, practices, health-seeking behaviours, and community capacity are the major factors influencing the behaviour change. Therefore, those were considered as the main components for behaviour change to achieve a sustainable dengue prevention in the highly endemic area in the district.

### The Modelling Phase

#### Step one

Overall goal was to decrease the morbidity and mortality due to dengue illness by improving the dengue prevention behaviours among the householders in highly endemic division in the district.

#### Step two

Expected preliminary behavioural objectives were formulated prior to develop Specific Behavioural Objectives (SBOO).

#### Step three

Current situational market analysis for communication keys was done using quantitative and qualitative methods. According to the quantitative analysis, mean knowledge on dengue prevention was 43.7% (SD 13.36; range 10 - 82). Of the participants, 57.7% (n=286) had good attitudes, 24.2% (n=120) had adequate dengue prevention practices, 44.6% (n=221) had good health seeking behaviours and 38.7% (n=74) had perceived that the community capacity is adequate for dengue prevention. Only 19% was knowledgeable on the importance of early notification of suspected cases and 19.2% (n=95) had adequate overall behaviours on dengue prevention. ‘Strengths, Weaknesses, Opportunities, Threats (SWOT)’ were assessed using qualitative method. The analysis of SWOT in relation to achieving the behavioural objectives are depicted in Table 1.

**Table 1.** SWOT analysis for dengue prevention behaviours.

|  |  |  |
| --- | --- | --- |
| **1.** | **Strengths** | Guidelines, monitoring and technical support from the National level (National Dengue Control Unit-NDCU)<br>Well established preventive sector with dedicated team for management<br>House to house inspection by Dengue Field Assistants at field level<br>School/Workplace dengue promotion activities<br>Support from other stakeholders<br>Availability of fogging facilities and case by case fogging activities |
| **2.** | **Weaknesses** | Delays in notification<br>Poor health seeking behaviours<br>Gaps in law enforcement<br>Lack of community participation<br>Not considered as a priority area prior to monsoonal period (lack in preparedness)<br>Poor attitudes of the community members<br>Improper urbanization<br>No proper monitoring and evaluation mechanism for dengue management activities<br>Poor waste management and no established waste management system<br>Lands without proper ownership<br>Gaps in national policies and legislations related to vector control<br>Inadequate financial allocation for dengue prevention activities |
| **3.** | **Opportunities** | Availability of Dengue Field Assistants at field level<br>Availability of fogging facilities<br>Waste collection by local government authorities<br>Dengue committee meetings at all levels (divisional level, district level and National level) with multi-stakeholder participation<br>National and regional level policies to prevent dengue<br>Opportunities and grants for research activities |
| **4.** | **Threats** | Political pressure for law enforcement<br>Increase in natural disasters which deviate the attention from dengue prevention to the disaster response activities |

Consumer need, cost, convenience analysis revealed that the majority (80%) is more benefit than the cost. Moreover, the people can earn extra money from manufacturing compost and home gardening using compost. Time taken for dengue activities is more beneficial than disease management. When considering the risk perception on dengue, it was very low and one-fourth of them perceived that it is not a deadly disease, and they believe so if their family is not affected by dengue. Moreover, when considering competitors, there were no alternative behaviours or services being offered for this community. But the priority is shifted to control other outbreak situations such as leishmaniasis control activities in the area. The preferred methods of communication among the community were group discussions (43.4%), awareness through telephones (17.3%), displaying of notices or handbills (15.7%), other modes like street drama (12%) and one to one communication (11.5%).

#### Step four

The Specific Behavioural Objectives (SBOO) were to adopt proper waste management practices according to 3R method (Reduce, Reuse and Re-cycling) among the householders to reduce vector density and to improve the regular weekly dengue prevention practices by allocating at least one day a week to practice 30 minutes of cleaning to improve dengue prevention behaviours. Branding the theme was planned to distribute among the participants of the workshops [Supplementary file 1].

#### Step five

The main strategy was to conduct a community empowerment programme to improve household waste management and weekly practices on dengue prevention by conducting ‘The administrative mobilization and public relationship, public relations, public advocacy, community mobilization, personal selling, advertising and promotion, and point of service promotion during follow up. Table 2 shows the activities which were identified by the developed COMBI based intervention.

**Table 2.** Activities to achieve targeted objectives to prevent dengue.

|  |  |
| --- | --- |
| **1.** | **Administrative mobilization and public relations** |
|  | To formulate a training manual of ‘COMBI based community empowering for sustainable dengue control’ for field officers with the support of public health specialists |
|  | • To conduct consultative meetings and Key Informant Interviews (KII) with health and non-health stakeholders to assess the need for dengue prevention in the area<br>• To discuss with the heads of healthcare institutions on how to prepare an action plan for dengue management for the healthcare institutions |
| **2.** | **Public Advocacy** |
|  | • To conduct advocacy programmes to motivate the officials of local government authority (“Pradeshiya Sabha”) of the area, religious leaders, and the community leaders to get the support for waste management |
| **3.** | **Community Mobilization** |
|  | • To identify volunteer leaders for the sustainability of the dengue prevention<br>• To train the leader group to conduct Training of Trainers (TOT) programme |
| **4.** | **Personal Selling, Advertising and Promotion** |
|  | • To select non-leader group and leader groups among the community<br>• To conduct training workshops for non-leader community participants with interactive discussion<br>• To brand the theme with the material |
| **5.** | **Point of service Promotion during follow up** |
|  | • To conduct weekly follow ups using a ‘Weekly follow up observation record’ in all three languages for four weeks |

Two-day workshops were planned to conduct for 25-30 participants per workshop. Day one [Session I] was a motivation and the awareness on dengue prevention, which was planned to conduct by the district Health Promotion Officer (HPO). Educational presentation on awareness on dengue and consequences of dengue disease, identification of dengue vectors and vector control strategies and the importance of at least 30 minutes premises inspection per week, waste management according to 3R concept, and interactive lecture on composting and organic farming were conducted during the session. Session II was a skill improvement session of two sub-components. Demonstration of composting, organic farming and interactive discussion session using an information guide with the participants and a session including constructive discussion with having feedback of peers and trainer through analyzing the problems of the workshop for the participants were planned to conduct as the component of session two. Day-two also has two sessions. A household inspection and entomology survey are planned to conduct by trained entomological assistants. During session II, practical demonstration of composting lead by the community leader group and community base small group discussion at the households to identify the practical problems. Thereafter, theoretical problems were planned to address.

#### Step six

The finalized plan was presented to the stakeholders to get their feedback prior to finalizing the intervention plan.

#### Step seven

The management team was consisted of the first author and public health specialists. The technical advisory group was consisted of public health specialists with the collaboration of Regional Director of Health services (RDHS), district HPO, and Provincial vector control officer, Medical Officer of Health (MOH) and, public health inspectors. The collaborating agencies were Ministry of Health, RDHS office, MOH office, Agriculture department and District secretariat, Kurunegala, Sri Lanka.

#### Step eight: Monitoring and Evaluation

Monitoring and evaluation were planned to conduct after the community empowerment program according to their needs. It was planned to follow up weekly for one month the research team following the implementation sessions. The follow up is mainly focused on personal selling and supplemented by the counselling after assessing their compliance. The desired behavioural objectives are planned to repeat weekly for four weeks, and post-intervention evaluation is planned to carry out after three months of completion of the intervention program. Weekly observational record was designed to document [Supplementary file 2].

#### Step nine: Impact Assessment

A pilot study was planned to conduct to assess the feasibility issues and outcomes of interest including changes in knowledge, attitudes, vector control practices, dengue prevention behaviour and community capacity. Entomological surveys were planned to carry out to assess the impact on vector densities. The behavioural impact was planned to assess by observations of management of waste according to 3R concept, outdoor, indoor water containers, water storages and roof gutter at the household level during the implementation of the finalized plan. After piloting the developed intervention package, a cluster randomized trial was planned to conduct to assess the effectiveness of the complex intervention package.

#### Step ten: Scheduling and Budget

The action plan was developed after taking expert opinion from the panel of expert.

## Discussion

The complex intervention was formulated according to the revised framework of complex intervention development. The rationale for a complex intervention is to develop a theoretical understanding of the likely process of change, by drawing on existing evidence and theory, supplemented, if necessary, by new primary research, qualitative approach with the stakeholders or the targeted population,[4]. In addition, there may be lots of competing or partly overlapping theories,[20]. However, the research team need expertise opinion on relevant disciplines to find the most appropriate theory and as the complex interventions have several dimensions of complexity, it may be to do with the range of possible outcomes, or their variability in the target population, rather than with the number of elements in the intervention package itself. It follows that there is no sharp boundary between simple and complex interventions, [4]. The interventions have been developed according to different models and planning processes. The planning of the intervention package was formulated using WHO’s 10 steps of COMBI planning method. The COMBI approach is successfully demonstrated that correct problem identification synergized with community engagement can potentially reduce Aedes proliferation and dengue morbidity in Malaysia and Sri Lanka,[21,22]. Out of the COMBI based interventions, Malaysia used integrated marketing communication techniques to inoculate this behavioral change to target group. Therefore, the COMBI approach is successful with correct problem identification and community engagement,[21]. The use of the Delphi technique instead of face-to-face consultative meetings had the advantage of not requiring the experts to take time off their schedules to contribute to the study. It allow the experts to respond at any time convenient to them and to contact any source of information if needed. Further, this process facilitated the independence of forming opinion and perspectives as it prevented the manipulation of opinion by influential individuals, which could happen in a face-to-face consultative meeting,[23]. In the process of development of the intervention plan, there was a quantitative study to finalize the SBOs. For that, validated tools were used to assess the current situation marketing analysis as a major step in the COMBI planning process. The generalizability of an intervention package depends on the effectiveness and validity of models, theories, methods use for development process. In the present study, after considering the effectiveness of the already conducted studies, planning abilities, implementation, and feasibility issues with the consensus of the expert panel, COMBI planning was identified for the development of the interventional package. Importantly, the COMBI theory was used to plan the process of community empowerment program, because it is a proven effective method for behavior changes for dengue control and widely utilizing planning process in different countries,[19]. Moreover, the process of assessing current situation market analysis also performed by mixed methodology with representative samples. Therefore, the generalizability of the intervention to high endemic areas could be done with adaptation to setting. Notably, incorporation of similar programs for prevention of dengue through behavioural change to the public health system of Sri Lanka seems feasible and cost effective,[22]. Moreover, a community-based dengue prevention and control process enables key stakeholders in the community to actively prevent and control their dengue problem, [18]. Importantly, the studies in the Dominican Republic,[24], Colombia,[25], Hulu Langat,[21] and Thailand,[18] revealed that the interpersonal communication was an effective way to achieve greater success of a community-based programme. Therefore, the present intervention package aimed to empower the community by developing a sustainable dengue control process at the grass root level with the involvement of multi-stakeholders. Importantly, the dengue problem can be solved by conducting the community-based dengue prevention process with active participation of key stakeholders in the community. Moreover, sustainability of a community-based dengue prevention and control comprise activities depends on the degree of eliminate larval breeding sources, control adult mosquitoes, apply personal protection, introduce dengue symptom detection, and outbreak prevention, [26]. In Sri Lanka also there is a clear need to address above areas especially waste management and vector control,[22]. Importantly, the specificity of the intervention messages of the present study was waste management according to 3R concept aiming source reduction from both outdoor and indoor vector breeding places. Formalization of waste management would empower the community members to identify breeding sites and removing them. It would not that difficult or not labour intensive or less costly than managing dengue cases or mass cleaning campaings. Such specific messages have been used to achieve successful behavioural outcomes in the interventions in other countries, [21,22,25,27,28]. Solid waste management is a growing challenge to many countries. Improper waste management serve as the breeding places for many vectors resulting in proliferation of vector-borne diseases,[29]. Moreover, the presence of solid waste around households, such as cans, car parts, bottles, old and used tyres, plastic materials, broken clay, glass vessels and coconut shells, created outdoor breeding sites for Aedes mosquitoes and represented in our ecosystem the most productive container types. Maintaining solid waste for a long time often more than seven days supports the breeding of Aedes aegypti,[30] and increases the transmission of dengue. Moreover, Wijesundara planned the intervention according to COMBI with two SBOs including 30 minutes’ inspection on Sunday and improve the proper waste management practices and Rozhan and others have identified the SBOs as management of water containers twice weekly and scrub any containers found to contain larvae. Furthermore, a study was carried out in Malaysia revealed that the knowledge, attitudes and practices were influenced by the interventional programme during its implementation weeks. However, the outcome evaluations at the end of the study revealed that COMBI programme failed to achieve the desired behavioral impact of the programme. The multi-stakeholder collaboration is one of the suggested solutions to overcome the problems of the programme, because there was lack of human resources and funding. Furthermore, they suggested to improve the health sector participation for the awareness of the community to prevent dengue,[31]. Therefore, for the development of the present intervention package, the multi-stakeholder participation was ensured. Similar to the present study, community empowerment programs were conducted in Cuba,[32,33] and Myanmar,[28]. Moreover, Behaviour change intervention package was developed to manage household water container in Philippine, 2012. Further, this study revealed that other factors such as social and political environment are needed to explain community responses to new dengue vector control interventions,[27]. Therefore, the present study was developed using new communication strategy with multi-stakeholder participation. Another study conducted in Cuba revealed that the importance of the community-based strategies such as organizational management, entomological risk surveillance, capacity building and community work for vector control. The community empowerment strategy increased community involvement and added effectiveness to routine *A. aegypti* control,[33]. The efficacy of community-controlled partnership-driven interventions was found to be superior to the vertical approach in terms of sustainability and community empowerment. Moreover, a study in Sri Lanka, revealed that the vector control interventions had a significant impact on vector densities (BI) and on dengue incidence. It revealed that rigorous vector control programs lead to reduce the disease and economic burden of dengue in endemic settings,[34] al, 2019). When considering the intervention packages in the studies, there was a lack of scientific evidence in the development process. Therefore, to develop the present intervention package, the evidence from most of the effective intervention packages were utilized. Not only that, but the identified gaps of the failed interventions were also considered to have better outcomes of interest. Therefore, the present intervention package can be considered as a scientifically developed intervention for sustainable dengue prevention.

## Conclusion and Recommendations

Developing a COMBI plan for an area after the identification of specific behavioural objectives would make it feasible for implementation in order to empower the community to prevent dengue in the area and to improve community health services. Scientifically developed COMBI based planning process can be used to bring about satisfactory control of dengue with the participation of the community in high endemic areas to achieve sustainable dengue prevention. This type of intervention can be applied to any locality after conducting the situation marketing analysis of the relevant area and developing area specific COMBI plan with the support of the preventive sector healthcare institutions in any country. Moreover, future research can be conducted using the COMBI planning process in the other endemic areas in preventing dengue outbreaks in the region or globally.

## Data Availability

All data produced in the present study are available upon reasonable request to the principal author

## Author contributions

Conceptualization and methodology: RMNUR, CA, AB; Technical advisors: NP, AA; Writing and Original draft preparation: RMNUR; Review, editing and supervision: CA, AB, NP, AA, SM.

## Footnote

### Conflicts of Interest

The authors declare no conflict of interest.

### Funding

This research did not receive any specific grant from funding agencies for publication.

### Institutional Review Board Statement

Ethical clearance was obtained from the ERC, Faculty of Medicine, University of Colombo, Sri Lanka (EC/18/134).

## Acknowledgements

- Provincial Directors of North-Western Province (NWP), Regional Directors of Kurunegala District, Consultant Community Physician, NWP, MOH Kurunegala, Public health team including Public health inspectors, dengue field assistance and all participants.
- Professor Everold Husein (Senior Communication Advisor-Consultant, Adjunct Professor, President-The COMBI Institute).

## Notes

### Competing Interest Statement

The authors have declared no competing interest.

### Funding Statement

This study was self-funded by the principal investigator

### Author Declarations

ERC of the University of Colombo Sri Lanka (EC/18/134)

### Summary of Updates

1. Abstract - Structured according to the JECH guideline 2. 2 Supplementary material included 3. Tables

