## Supplementary file 1 for "DEVELOPMENT OF A COMPLEX INTERVENTION PACKAGE FOR DENGUE PREVENTION"

Supplementary file 1 : Theme of the behavioural change

**Theme of the Behavioral Change:**

### Waste Management

**According to**

### 3R - Concept

### (R-REDUCE, R-REUSE, R-RECYCLE)

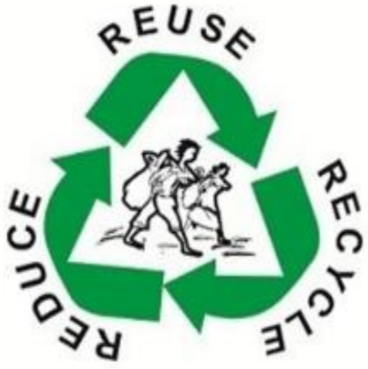


### Clean Your Premises

### At least 30 minutes Per a Week

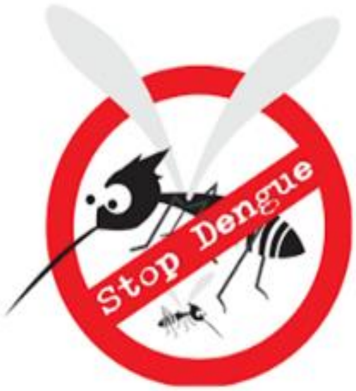


### Let's Have a Dengue Free Environment
