## Supplementary file 2 for "DEVELOPMENT OF A COMPLEX INTERVENTION PACKAGE FOR DENGUE PREVENTION"

The Weekly Record for Observation of Dengue Mosquito Breeding Places

In this week.....

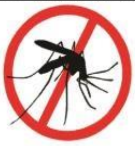

Shall we remove vector breeding places?

Your honest response is greatly appreciated!!!!

If you perform it mark a 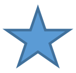 in the relevant day...

| Month | Week | Monday | Tuesday | Wednesday | Thursday | Friday | Saturday | Sunday |
|---|---|---|---|---|---|---|---|---|
| 1 | 1 | | | | | | | |
| | 2 | | | | | | | |
| | 3 | | | | | | | |
| | 4 | | | | | | | |
| 2 | 1 | | | | | | | |
| | 2 | | | | | | | |
| | 3 | | | | | | | |
| | 4 | | | | | | | |
| 3 | 1 | | | | | | | |
| | 2 | | | | | | | |
| | 3 | | | | | | | |
| | 4 | | | | | | | |

Remarks: (If any)

..................................................................................................................................................
..................................................................................................................................................
..................................................................................................................................................

Please keep this form safely and handover to the dengue research team members who visited to your place. Your regular good practice will save a life

[For Further clarification; please do not hesitate to contact the principal investigator

Dr. Nayani Rajapaksha +94778974176]

*For official use only* **PHI area:**

**Cluster number:**

**Serial number of the household:**
